# Genetic Markers and Predictive Factors Influencing the Aggressive Behavior of Cerebral Cavernous Malformation

**DOI:** 10.1101/2023.12.30.23300669

**Authors:** Gustavo F. Galvão, Luisa M. Trefilio, Andreza L. Salvio, Elielson V. da Silva, Soniza V. Alves-Leon, Fabrícia L. Fontes-Dantas, Jorge M. de Souza

## Abstract

Biological behavior of Cerebral Cavernous Malformation (CCM) is still controversial without clear-cut signature for biological mechanistic explanation of lesion aggressiveness. There is plenty evidence implicating dysregulated inflammatory and immune responses in vascular malformation pathogenesis, including CCM. In the present study, we evaluated the predictive capacity of the SNPs *VDR^rs7975232^, VDR^rs731236^, VDR*^rs11568820^ as well as expanded the analysis of *PTPN2*^rs72872125^ and *FCGR2A^rs1801274^* in relation to the aggressive behavior of CCM and its implications in biological processes. This was a single-site prospective observational cohort study with 103 patients enrolled, 42 had close follow-up visits for a period of 4 years, focused on 2 main aspects of the disease: (1) symptomatic event that composed both intracranial bleeding or epilepsy and (2) precocity of symptoms. We report a novel observation that the *PTPN2*^rs72872125^ CT and the *VDR*^rs7975232^ CC genotype were independently associated with an asymptomatic phenotype. Additionally, *PTPN2*^rs72872125^ CC genotype and serum level of GM-CSF could predict a diagnostic association with symptomatic phenotype in CCM patients, while the *FCGR2A^rs1801274^* GG genotype could predict a symptomatic event during follow-up. The study also found a correlation between *VDR^rs731236^* AA and *VDR*^rs11568820^ CC genotype to the time to first symptomatic event. In summary, this study provides valuable insights into the genetic markers that could potentially impact the development and advancement of CCM.

## INTRODUCTION

Cerebral cavernous malformation (CCM) are vascular malformations consisting of capillary-like channels with a single layer of endothelium and no intervening nervous tissue^1,2^. CCM are among the most common vascular malformation of the central nervous system (CNS) affecting 0.5%–1% of the population, with magnetic resonance image (MRI) as the gold standard for the diagnosis and the lesions evaluation should include the hemosiderin-sensitive techniques as the susceptibility-weighted imaging (SWI)^3,4^. Intracerebral hemorrhage (ICH) and seizure, are the main clinical manifestations, followed by neurological disability without evidence of bleeding. However, many patients remain asymptomatic throughout their life and it is not known what factors may predict aggressive manifestations in individual cases^4,5^.

Usually, single cavernomas are detected in patients affected by sporadic forms and often associated with a developmental venous anomaly. Familial CCM is mostly associated with the occurrence of multiple lesions that might increase in number and size with aging ^6^. The familial form of the disease is linked to mutations on specific genes, (*CCM1/KRIT1*, *CCM2/MGC4607* and *CCM3/PDCD10*), which encode distinct proteins involved in the endothelial cell junction function and in the interaction with cytoskeletal proteins. The most frequent and well-studied of the CCM genes is the *CCM1/KRIT1*, with mutational analyses showing a Hispanic-American ancestral haplotype^7,8^. CCMs are characterized by an incomplete disease penetrance, being 80% to the CCM1 form, near 100% to the CCM2 and 60% to the CCM3 form^9,10^. Furthermore, the discovery of *PIK3CA* mutations in CCM lesions provides important new insights into the genetic factors that contribute to the variable expression and progression of the disease phenotype ^11^.

Biological behavior of CCM is still controversial without clear-cut signature for biological mechanistic explanation of lesion aggressiveness. There is plenty evidence implicating dysregulated inflammatory and immune responses in vascular malformation pathogenesis, including CCM. Several dysregulated pathways have been confirmed in the transcriptome of CCM lesions, which are out of balance having an impact on the permeability of the blood-brain barrier and the stability of endothelial tight junctions ^12^. Notably, inflammatory and immune cells such as monocytes, macrophages, B and T cells, are present in human CCM lesions as well as in mouse models of CCM^13–17^. Recently, our group have demonstrated an adaptive immune-cellular reaction to CCM within CD20+ and CD68+ in the pericavernous tissue of an aggressive pharmaco-resistant epilepsy patient^18^. Moreover, the severity of the disease has been linked to genetic polymorphisms within inflammatory and immune response genes ^19^.

The significance of certain genetic variants in the development of CCM has been established, underscoring the need for further investigation into their specific roles. Tang et al 2017 showed allele T for SNP in *TLR4* (rs10759930) gene is associated with increased CCM lesion number ^20^. Some studies have also identified genetic polymorphisms in genes related to oxidative stress with a significant impact on inter-individual variability in CCM disease onset and severity^21^. Of particular interest, in a previous study, our group demonstrated the association of *FCGR2A^rs1801274^* GG and *PTPN2*^rs72872125^ CT genotype with a symptomatic profile of CCM patients in a smaller cohort ^22^.

In the present study, we evaluated the predictive capacity of the SNPs *VDR^rs7975232^, VDR^rs731236^, VDR*^rs11568820^ as well as expanded the analysis of *PTPN2*^rs72872125^ and *FCGR2A^rs1801274^* in relation to the aggressive behavior of CCM and its implications in biological processes. We also looked for plasmatic inflammatory cytokines expressed in the patients, verifying a pattern of heterogeneity of plasmatic expression and any correlation with the genetic variation identified with different clinical phenotypes of CCM. Using a multi-step Bayesian approach, we thought to build a biomarker that could predict a diagnostic and a prognostic aggressive phenotype of CCM.

## METHODS

### Study Design and Population

This was a single-site prospective observational cohort study. Qualified researcher on human subjects conducted this study having been approved by the National Council for Ethics in Research (CAAE 69409617.9.0000.5258). Blood samples were collected between October 2017 to March 2023 and informed consent was obtained. Strobe criteria were used to report the findings of this study ^63^.

The epidemiological data were defined at each clinical assessment. Patients were stratified as multiform when they harbor multiples CCM on SWI or Gecho or as isolated form when they had a single lesion at the CNS. Both definitions were based on MRI study with SWI or Gecho and the absence of venous development anomalies in the multiple cases in proximity.

CCM clinical presentation were identified as symptomatic (hemorrhage or seizure) and asymptomatic. Intracerebral hemorrhage was defined based on the consensus of Cavernous Angiomas with Symptomatic Hemorrhage (CASH) ^50^. The solely epileptic patients were defined as not having achieved previous hemorrhagic consensus and whom the MRI evaluation depicted no hint of hemosiderin beyond the usual smooth and regular deposit around a CCM lesion.

In this study, we focused in 2 mains aspects of the disease: (1) symptomatic event that composed both intracranial bleeding or epilepsy and (2) precocity of symptoms. We considered that these presentations may reflect an aggressive phenotype of the disease. We also designed six sub-group analysis in order to check for correlation - 60 years or older, female, familiar form, pure epileptic without signs of lesion bleeding, pure symptomatic bleeding and infratentorial lesion. Among the 103 consecutive cases, 42 had closely follow-up visit for a period of 4 years. The remaining patients either did not have a clinical follow-up visit or underwent surgical resection.

### Sample collection and processing

Blood samples were collected with EDTA and processed by centrifugation at 720 × g and 4 °C for 5 min to separate the plasma. The plasma supernatants were immediately stored at -80°C until further processing. The Genomic DNA (gDNA) extraction was performed using the PureLink Genomic DNA Mini Kit according to the manufacturer’s recommendations (ThermoFisher Scientific, Waltham, Massachusetts, USA). The quality of gDNA was determined by NanoDrop 2000 (ThermoFisher Scientific) followed by quantification using the Qubit dsDNA HS Assay Kit (ThermoFisher Scientific) and Qubit Fluorometer3.0 (Thermo Fisher Scientific).

### Plasma isolation and Inflammatory Modulators Assessment

Proinflammatory cytokine and chemokine levels in plasma were measured using a multiparametric immunoassay based on XMap-labeled magnetic microbeads (Luminex Corp – Austin, TX, USA). A human ProcartaPlex ™ Panel (Invitrogen – Waltham, MA, USA) was used to analyze a set of 18 cytokines and chemokines (IL-1β, IL-2, IL-4, IL-5, IL-6, IL-9, IL-10, IL-12p70, IL-13, IL-17A, IL-18, IL-21, IL-22, IL-23, IL-27, GMCSF and INF-γ). The samples were measured according to the manufacturer’s instructions and simultaneously to avoid potential batch, as described previously, using a BioPlex MAGPIX system (Biorad -Hercules, CA, USA) ^64,65^. Cyto/chemokine expression was measured in duplicate, and the levels of inflammatory modulators were analyzed using Xponent v. 3.0 software (Luminexcorp) and expressed in pg/ml.

### Analysis of polymorphism of PTPN2^rs72872125^, VDR^rs7975232^, VDR^rs731236^, VDR^rs11568820^ and FCGR2A^rs1801274^ genes

The SNPs investigated in this study are detailed in Table 1. The SNPs were genotyped by allelic discrimination performed in QuantStudio™ 3 Real-Time PCR System (ThermoFisher) using TaqMan SNP genotyping assays (ThermoFisher). The characteristics of the *PTPN2*^rs72872125^, *VDR^rs7975232^, VDR^rs731236^, VDR*^rs11568820^ *and FCGR2A^rs1801274^* genes were obtained from the SNP bank of the National Center of Biotechnology Information - NCBI (http://www.ncbi.nlm.nih.gov/). The probes for each SNP were produced by Applied Biosystems ™ rs72872125 (C 98019281_10), rs7975232 (C 28977635_10), rs731236 (C 2404008_10), rs11568820 (C 2880808_10) and rs1801274 (9077561_20). As the assay was designed and standardized to work with the same thermal cycles, a single protocol was applied for both genes. Briefly, PCR was performed with a 25 uL reaction mixture containing 10 ng DNA, TaqMan®_Universal PCR Master Mix (1X), Probe TaqMan® _Gene Expression Assay (1X), and DNAse free water for the final volume. The Real Time PCR conditions were: initially 60°C for 30 seconds and then 95°C for 10 min, and subsequently 40 cycles of amplification (95 °C for 15 seconds and 60°C for 1 min), and then 60°C for 30 seconds. The five selected SNPs were amplified on separate plates.

**Table 1:** Studied SNPs and their basic characteristics and female sex adjusted multivariable analysis.

| Gene | SNP ID | Alternative<br>Name | Chr/Location | Nucleotide<br>Change | Aminoacid<br>Change | MAF<br>ALFA |
| --- | --- | --- | --- | --- | --- | --- |
| *PTPN2* | rs72872125 |  | 18/intron | C>T |  | 0.11 |
| *VDR* | rs7975232 | Apa1 | 12/intron | C>A |  | 0.55 |
|  | rs731236 | Taq1 | 12/exon | A>G | Ile > Ile | 0.38 |
|  | rs11568820 |  | 12/intron | C>T |  | 0.28 |
| *FCGR2A* | rs1801274 |  | 1/exon | A>G | His > Arg | 0.51 |

Chr - chromosome; MAF - minimal allele frequency

### In silico Structural Prediction of FCGR2A^rs1801274^ Protein and Phylogenetic Analysis

Since the *FCGR2A^rs1801274^* SNP is the only with amino acid change, we performed a structural analysis using bioinformatic tools. *FCGR2A^rs1801274^* transcript structure was predict by directly modifications of the wild-type *FCGR2A* using FASTA sequence obtained on NCBI database ^66^. The new FASTA file was submitted to the Chimera software (developed by the Resource for Biocomputing, Visualization, and Informatics at the University of California, San Francisco, with support from NIH P41-GM103311) for overall structural analysis ^67^.

The evolutionary history was inferred using the MEGA-X software using the maximum likelihood method and Jones-Taylor-Thornton (JTT) matrix-based model. We first gathered the *FCGR2A* wild-type sequence from Homo sapiens and 19 other mammals to study the evolutionary conservation of this protein. Structural alignment was performed using the muscle algorithm that allows multiple sequencing alignment with high accuracy and high throughput ^68^.

### Statistical Analysis

Statistical analyses were performed with STATA 13 (StataCorp LP, TX, USA) and GraphPad Prism 9.0.0 MacOS (GraphPad Software, San Diego, California USA, www.graphpad.com). Categorical variables were expressed as n (%) and continuous data were given as mean and standard deviation (SD). A 2-tailed 2-sample t test or Mann-Whitney test was applied for continuous variables while the association between two categorical variables was measured by Pearson Chi-square (χ2) test when appropriate.

Allelic and genotypic frequencies were calculated for patient and control subjects via direct gene counting. Genotypic distributions in Hardy–Weinberg equilibrium (HWE) were evaluated by two-tailed χ2-test. Linkage disequilibrium (LD) were analyzed using: Linkage Disequilibrium Calculator (https://grch37.ensembl.org/Homo_sapiens/Tools/LD), LDlink on https://ldlink.nih.gov/ and Heatmap was plotted by https://www.bioinformatics.com.cn/en.The degree of LD between SNPs is represented by R^2^ and D’.

Genetic polymorphisms were evaluated through a univariate and multivariable binary logistic regression adjusting for sex, age and familiar form when adequate in matter of phenotype (symptomatic and asymptomatic). Plasma cytokines were also evaluated between genotypes and phenotypes (symptomatic and asymptomatic). Values over 2 times de standard deviation were excluded from analysis due to possible bias results.

Genetic polymorphism and plasma cytokines verified as statistically different between groups (both diagnostic and prognostic) were combined through a canonical discriminant function analysis in order to build a biomarker-model of symptomatic profile. All the possible combinations showing significant association in symptomatic individuals were build. Receiver Operating Characteristic (ROC) curves were generated along with a computed area under the curve (AUC) for each combination individually and ROC curves were compared to identify if any of them were statically superior then the others. The best model-biomarker to differentiate symptomatic and asymptomatic patients in both diagnostic and prognostic scenario was selected according to the Akaike Information Criteria (AIC), representing the best fit parsimonious model to the data with the fewest number of predictors. The optimal cutoff point was generated from ROC curves utilizing the Youden index method. Mann-Whitney test was then used to verify the difference in values generated using the modeled equation among symptomatic and asymptomatic patients and a logistic regression was used to control for age and sex. A p value <0.05 was considered significant.

To balance baseline covariates between patients with and without symptoms during follow-up we used a propensity score matching (PSM) strategy. The strategy involved 1:1 pairing and nearest-neighbor methods. After PSM, the distribution of gender, familial form, and age was balanced between the groups and the biomarker analysis was remade.

Finally, A Kaplan-Meyer survivor analysis was carried out among patients who presented different polymorphism combinations. Hazard ratios were calculated with 95% confidence intervals through a log-rank test. Failure events were determined according to if there had been symptomatic presentation at some point in the patient’s lifetime

## Results

### Demographic and CCM lesions characteristics

Out of 103 CCM patients enrolled in the study, 44 are multifocal/familial CCMs, 70 patients presented with symptomatic phenotype, which 48 presented a symptomatic hemorrhage secondary to their CCM, and 22 presented with seizures and no history or signs of CCM bleeding on MRI. The median age of enrolment was 45.6, within a mean age of 41.6 in the symptomatic group and 53.7 in epileptic sub-group. There were no significant differences in terms of sex, age and form (sporadic vs familiar/multifocal) between the outcome groups (Table 2).

**Table 2:** Summary of the Demographic and Clinical Characteristics.

| Characteristics | N (%) | Multifocal/Familiar | Age (years), mean | Sex: Female, n |
| --- | --- | --- | --- | --- |
| Asymptomatic | 33 (32%) | 13 | 46.3 | 22 |
| Symptomatic | 70 (67%) | 31 | 45.3 | 43 |
| Hemorrhage | 48 (46%) | 20 | 41.6 | 33 |
| Epilepsy | 22 (21%) | 11 | 53.7 | 10 |
| Total | 103 | 44 | 45.6 | 65 |

N - absolute number

### Association of PTPN2^rs72872125^ and VDR^rs7975232^ to a Symptomatic Phenotype and Plasmatic Cytokine Levels

The distribution of the genotypic frequencies of the *PTPN2*^rs72872125^, *VDR^rs7975232^, VDR^rs731236^, VDR*^rs11568820^ *and FCGR2A^rs1801274^* in symptomatic and asymptomatic patients are shown in Table 3. All genotypic distributions were in Hardy-Weinberg equilibrium. Haplotype analysis indicated linkage disequilibrium between *VDR^rs7975232^* and *VDR^rs731236^* (D’>0.99), corroborating what has already been found in other populations (Figure 1A and Supplementary material). While the other combinations segregate independently *VDR*^rs11568820^ and *VDR*^rs731236^ (D’>0.04), *VDR*^rs11568820^ and *VDR ^rs7975232^* (D’>0.317) (Figure 1A and Supplementary material). A higher frequency of the *PTPN2*^rs72872125^ CT genotype (OR 0.34, 95% CI 0.11-0.99, p = 0.04) and the *VDR^rs7975232^* CC genotype (OR 0.06, 95% CI 0.006-0.612, p = 0.017) was observed in asymptomatic phenotype in the age, familiar and female adjusted multivariable analysis, when compared with symptomatic patients (Table 3). The ROC analysis (Figure 1B) revealed that patients with *PTPN2*^rs72872125^ CT genotype had a modest Area Under de Curve (AUC 0.420, SE 0.05 CI 95% 0.334-0.506) and *VDR^rs7975232^* CC individuals (Figure 1C) had a pour accuracy (AUC 0.439, SE 0.03 CI 95% 0.370-0.508).

**Figure 1:**
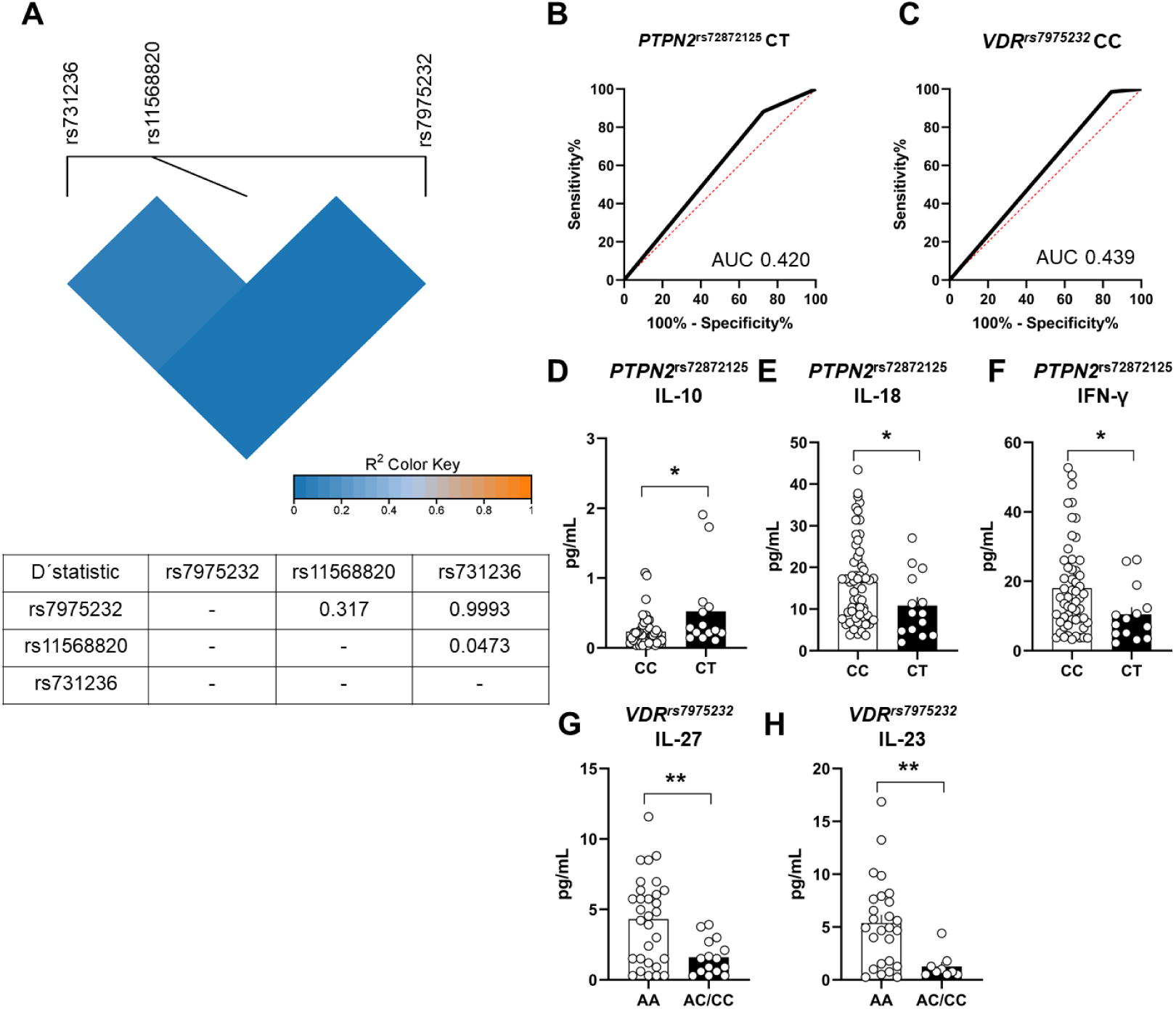
Association of PTPN2 and VDR to a Symptomatic Phenotype, Linkage Disequilibrium Map and Plasmatic Cytokine Expression. (A) Haplotype map showing the 3 SNPs analyzed and the range of R^2^ value, suggesting that VDR^rs7975232^ and VDR^rs731236^ are in linkage disequilibrium (D’>0.99), while the other combinations segregate independently VDR^rs11568820^ and VDR^rs731236^ (D’>0.04), VDR^rs11568820^ and VDR ^rs7975232^ (D’>0.317). (B) ROC analysis evidencing the performance of PTPN2^rs72872125^ CT genotype (AUC 0.420) and (C) VDR^rs7975232^ CC (AUC 0.439) to distinguish symptomatic patients. (D) Plasmatic cytokine expression between PTPN2^rs72872125^ groups showing the PTPN2^rs72872125^ CT genotype have higher plasma level of IL-10 (p = 0.0146), (E) low levels of IL-18 (p = 0.0450) and (F) low levels of IFN-γ (p = 0.0310) (Figure 1F), while individuals with at least one VDR^rs7975232^ C allele had (G) low plasma levels of IL-27 (p = 0.0055) and (H) low plasma levels of IL-23 (p =0.0034).

**Table 3:**
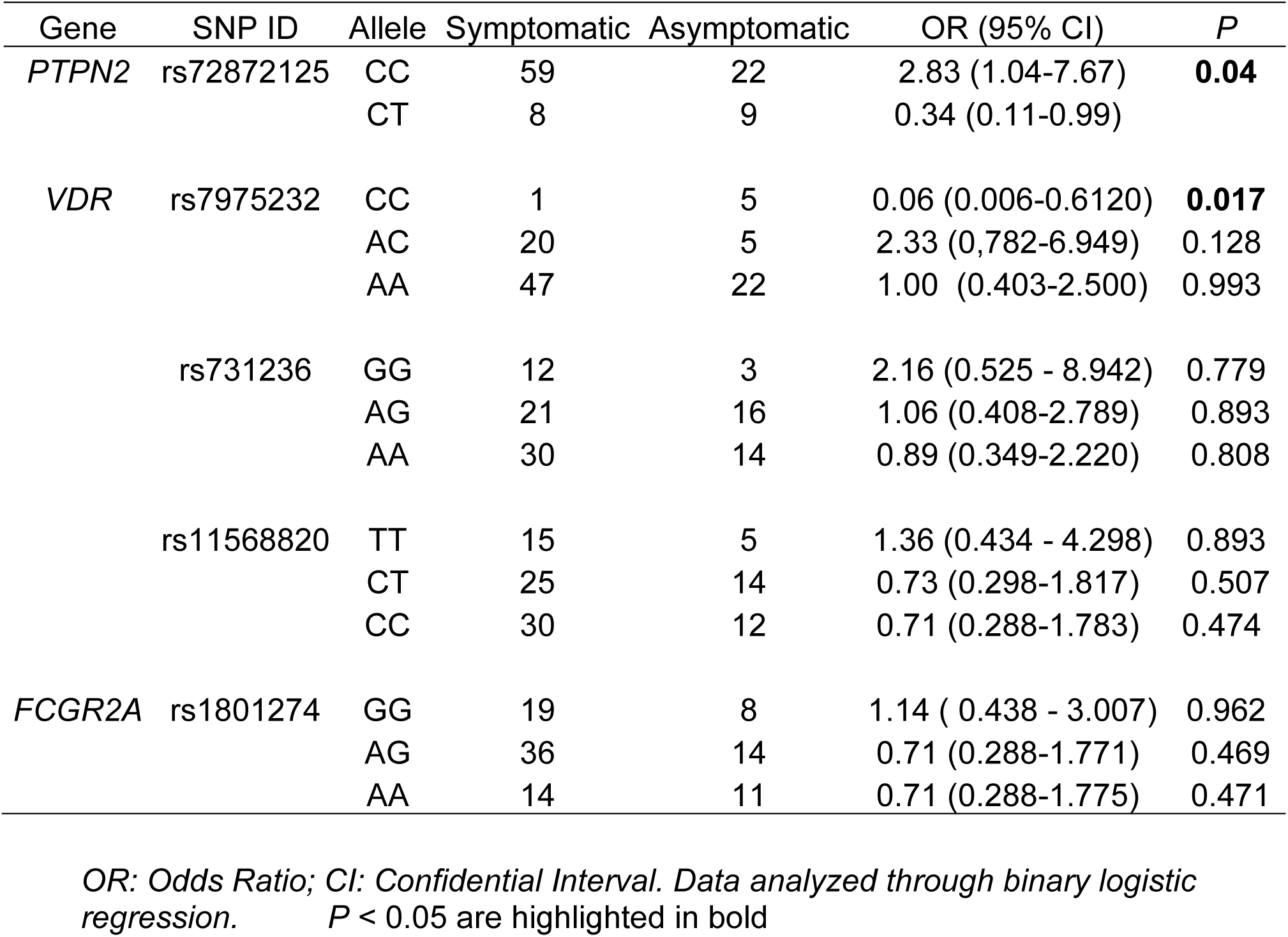
Association between SNPs and Different Phenotypes in the age, familiar and female sex adjusted multivariable analysis.

We also hypothesized that these genetic variants might influence the plasmatic inflammation signature of the CCM patients. We found that patients who harbor the *PTPN2*^rs72872125^ CT genotype showed a higher plasma level of IL-10 (*p* = 0.0146) (Figure 1D) and low levels of IL-18 (*p* = 0.0450) (Figure 1E) and IFN-γ (*p* = 0.0310) (Figure 1F), while individuals with *VDR^rs7975232^ at least one* C allele had low plasma levels of IL-27 (*p* = 0.0055) and IL-23 (*p* =0.0034) (Figure 1G-H). For other SNPs no significant difference in the levels of immune markers was observed. We did not observe any significant differences in the levels of these plasma molecules in relation to patient sex and form (sporadic/solitary or familial/multifocal).

### Performance of Diagnostic Biomarker

In order to build a diagnostic biomarker of CCM activity that could improve our previously published weighted biomarker formula, we tested new combinations of genetic and cytokines factors. First, the previously published diagnostic weighted biomarker was again confirmed to distinguish symptomatic and asymptomatic patients with 14.2% sensitivity and 97.4% specificity (AUC 0.641 SE 0.06 CI 95% 0.512 – 0.770, p =0.03), worse than the range reported previously. A similar canonical discriminant analysis approach was then implemented to determine if a weighted combination of different SNPs and cytokines could improve the diagnostic association with symptomatic phenotype. The best weighted-biomarker included the *PTPN2*^rs72872125^ CC genotype and GMCSF plasma levels as formulated -0,89*(GMCSF)+0,41*(*PTPN2*^rs72872125^CC) (AUC 0.663, SE 0.06 CI 95% 0.534 – 0.792, p = 0.01) with a specificity and sensitivity 85.7% and 41.3%, respectively (Figure 2A). This formula outperformed all the other possible formula (Table 1 Supplement) that had a statistically significant value. The median weighted combination value was 2.65-times increased (*p* =0.01) in symptomatic patients (median estimated value -0.36) compared with asymptomatic individuals (median estimated value -0.97) (Figure 2B). The formula performed fairly in the sub-group analysis of hemorrhagic patients (AUC 0.661, SE 0.07, CI 95% 0.517 – 0.806) but had a good accuracy in familiar form of the disease (AUC 0.774, SE 0.09, CI 95% 0.595 – 0.953, p = 0.01) and patients with infratentorial lesions (AUC 0.804, SE 0.08, CI 95% 0.642 – 0.966, p = 0.001) (Figure 2 C-D). Female symptomatic patients, pure epileptic patients and elderly patients were not statically significant.

**Figure 2:**
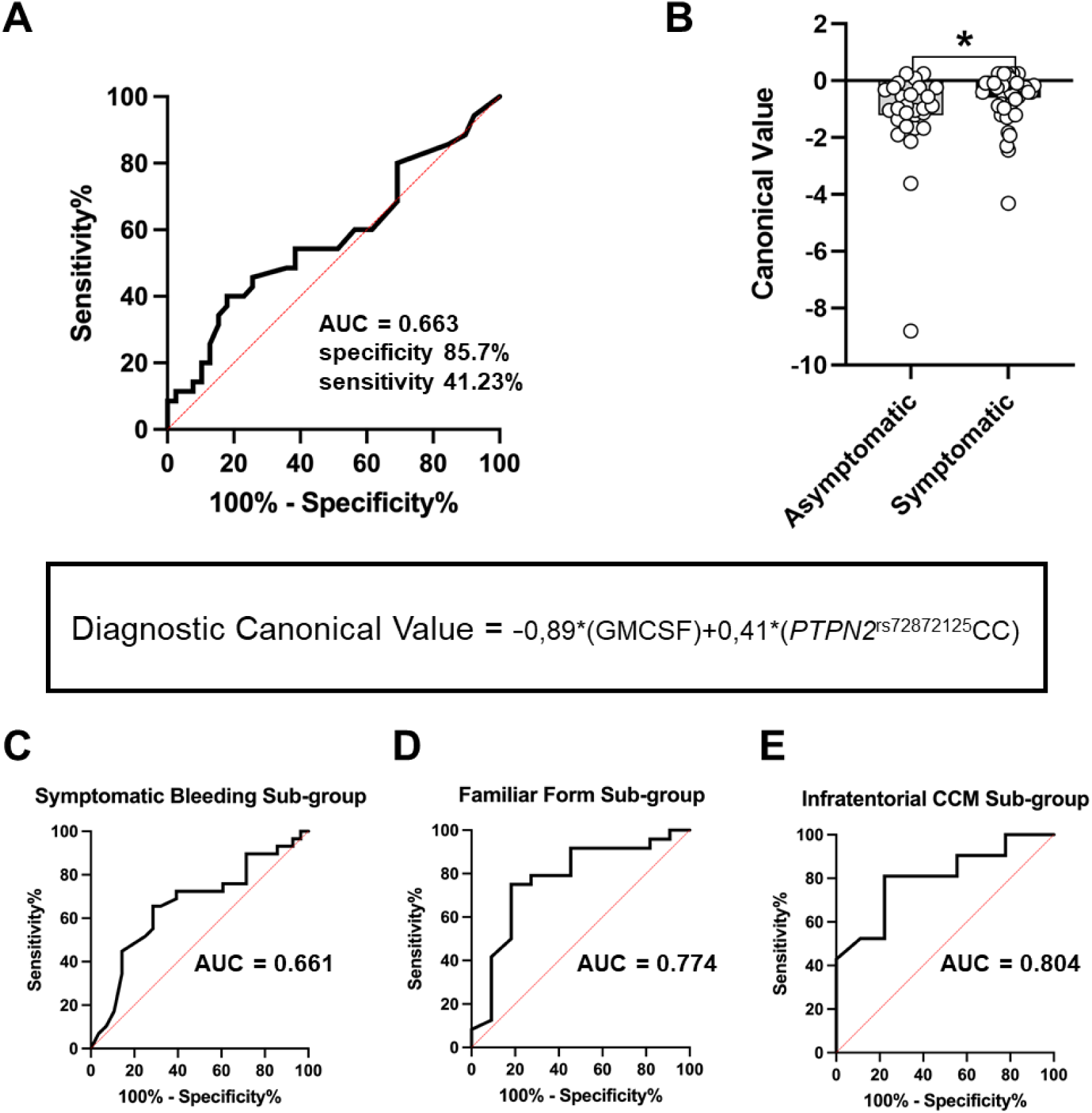
*Performance of Diagnostic Biomarker.* (A) ROC curve of the best weighted-biomarker included *PTPN2*^rs72872125^ CC genotype and GMCSF plasma levels as formulated -0,89*(GMCSF)+0,41*(*PTPN2*^rs72872125^CC) (AUC 0.663, SE 0.06 CI 95% 0.534 – 0.792, p = 0.01) with a specificity and sensitivity 85.7% and 41.3%, respectively. (B) The median weighted combination value was 2.65-times increased (*p* =0.01) in symptomatic patients (median estimated value -0.36) compared with asymptomatic individuals (median estimated value -0.97). Sub-group analysis demonstrating that the formula performed fairly in the sub-group analysis of (C) symptomatic bleeding patients (AUC 0.661, SE 0.07, CI 95% 0.517 – 0.806) but had a good accuracy in (D) familiar form of the disease (AUC 0.774, SE 0.09, CI 95% 0.595 – 0.953, p = 0.01) and (E) patients with infratentorial lesions (AUC 0.804, SE 0.08, CI 95% 0.642 – 0.966, p = 0.001).

### Validation Cohort with Symptomatic Event during Follow-up: Building a Prognostic Biomarker

We then tested if the newly weighted-diagnostic biomarker could act as a prognostic biomarker in a sub-group analysis of 42 patients that were prospectively followed-up after initial blood collection. 4 patients experienced a symptomatic event (1 lesion growth, 2 symptomatic hemorrhage and 1 newly onset epilepsy without signs of bleeding) while the others 38 stayed asymptomatic during the next 4 years. The weighted-biomarker had a fair accuracy (AUC 0.644, SE 0.078, CI 95% 0.491-0.976) in distinguishing these patients (Figure 3A). In order to verify if other combination of genetic variants and cytokines could act as a prognostic biomarker, we first tested the 5 SNPs in the sub-group of 42 patients that were closely followed-up. We found an independent statistically significant association of the *FCGR2A^rs1801274^* GG genotype (OR 16.10, 95% CI 1.32 – 195.52, p = 0.029) in the age and familiar adjusted multivariable analysis. This variant had an excellent accuracy (AUC 0.796, SE 0.12, CI 95% 0.631-0.897) in distinguishing these individuals (Figure 3B). We further tested the same gene in a propensity matched subgroup. The presence of the *FCGR2A^rs1801274^* GG genotype could predict a symptomatic event with high accuracy (AUC 0.875, SE 0.12, CI 95% 0.473-0.996) (Figure 3C). We also tested if this mutation would maintain its accuracy power when comparing symptomatic patients during follow-up and individuals who have never experienced any symptoms. We also found that *FCGR2A^rs1801274^* GG genotype could predict a symptomatic event with high accuracy (AUC 0.829, SE 0.14, CI 95% 0.519 - 0.956) (Figure 3D).

**Figure 3:**
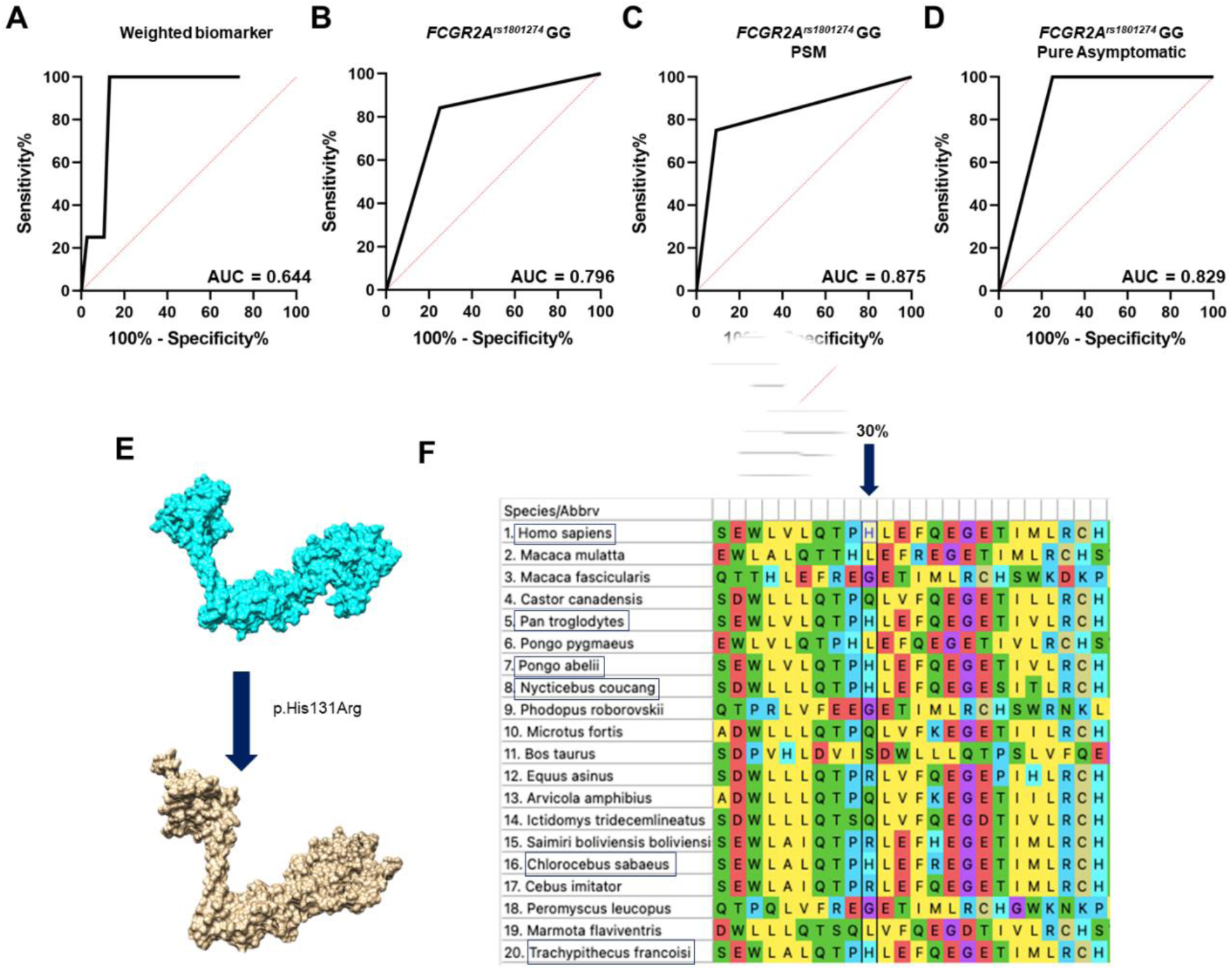
*Performance of Prognostic Biomarker and In Silico Structural Analysis of Transcript FCGR2A^rs180127^*. (A) The weighted-biomarker had a fair accuracy (AUC 0.644, SE 0.078, CI 95% 0.491-0.976) as prognostic biomarker while *FCGR2A^rs1801274^* GG (B) genotype had an excellent accuracy (AUC 0.796, SE 0.12, CI 95% 0.631-0.897) in distinguishing these patients. (C) ROC curve evidencing the performance of the *FCGR2A^rs1801274^* GG genotype after PSM analysis (AUC 0.875) and (D) in a sub-group of pure asymptomatic patients (AUC 0.829). (E) *In silico* structural analysis prediction reveals that the *FCGR2A^rs1801274^* leads to a substitution of a histidine for an arginine at position 131 leading to a predicted conformational structure slightly different than the wild-type. (F) Structural alignment predicts that the mutation c.500A>G (p.His131Arg) occurred in domain conserved in higher primates, within 30% of the species sharing this site.

### In Silico Structural Analysis of Transcript FCGR2A^rs1801274^

*In silico* structural analysis prediction reveals that the *FCGR2A^rs1801274^* leads to a substitution of a histidine for an arginine at position 131 (Figure 3E). The structural alignment predicts that the variation c.500A>G (p.His131Arg) occurred in domain that appears to be conserved mainly in higher primates, within 30% of the species sharing this site (Figure 3F).

### VDR variants relates to Precocity of symptoms

We tested if any of the *VDR* genetic variants could relates to a precocity of symptoms in the CCM cohort. Patients with the *VDR^rs731236^* AA genotype tend to present their symptoms late in their life compared to those who did not have (HR 0.52, SE 0.14, CI 95% 0.299-0.909 *p* = 0.020) (Figure 4A). Also, individuals that harbor the *VDR^rs^*^11568820^ CC genotype tend to have symptoms earlier (HR 1.58, SE 0.46, CI 95% 1.092-2.282 *p* = 0.016 (Figure 3B).

**Figure 4:**
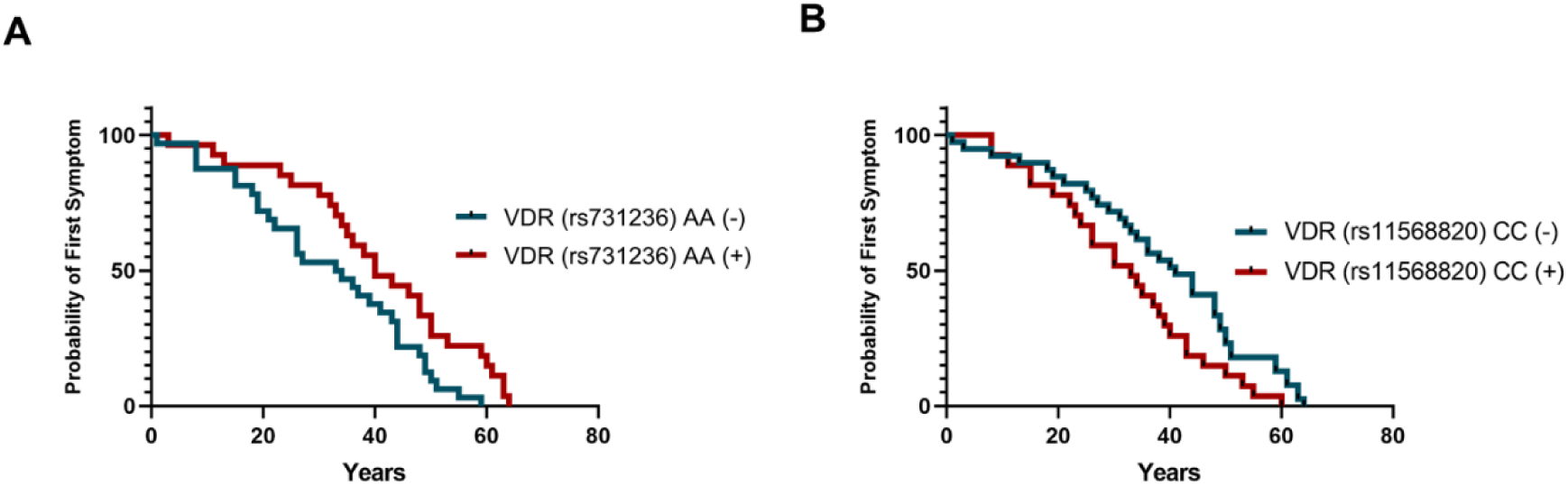
Kaplan-Meier curve demonstrating that VDR variants are related to Precocity of symptoms. (A) *VDR^rs731236^* AA genotype tend to have symptoms later in life compared to those who did not have (HR 0.52, *p* = 0.020). (B) Individuals that harbor the *VDR^rs^*^11568820^ CC genotype tend to have symptoms earlier (HR 1.58, *p* = 0.016).

## Discussion

Neuroinflammation is increasingly a focus of research in symptomatic events in CCM patients ^23^. In the present study, we evaluated a large and phenotypically well characterized CCM cohort and provided some key information about the genetic influence on the behavior of this unique neurovascular disease. Here, we have found an individual higher frequency of *PTPN2*^rs72872125^ CT and the *VDR*^rs7975232^ CC genotypes in asymptomatic patients, associated with changes in cytokine levels, suggesting a possible protective role. Our main new observation is that the combination of a balanced formula using the *PTPN2*^rs72872125^ CC genotype and serum level of GM-CSF could predict a diagnostic association with symptomatic phenotype in CCM patients, while the *FCGR2A^rs1801274^* GG genotype showed the best accuracy in predicting a symptomatic event in the next years, possibly functioning as prognostic genetic biomarker. In previous studies, we reported the association of this same variants with an aggressive phenotype of cerebral cavernous malformation but in a smaller sample^22^. Furthermore, another exciting discovery is that individuals with the *VDR^rs731236^* AA and *VDR^rs^*^11568820^ CC genotypes may experience an earlier onset of symptoms in the CCM cohort.

To date, there have been few published studies on SNPs in the *PTPN2* gene, and among them, a weak association with certain diseases or different outcomes was found. However, it is important to highlight that in combination with other variants they have been shown to possibly increase the susceptibility of chronic inflammatory disorders, including rheumatoid arthritis, type 1 diabetes and celiac disease^24–28^. Furthermore, there have been research findings indicating the presence of epistasis between *PTPN2* and *VDR* gene ^29,30^. Interestingly, patients who inherit the *PTPN2*^rs72872125^ CT genotype have high levels of IL-10 when compared with CC genotype (Figure 1). IL-10 is an anti-inflammatory molecule, with a well-established function in restraining and regulating both acute and chronic inflammatory processes ^3132^. Lyne et al. (2019) demonstrated that IL-10 is an important molecule present in diagnostic CASH biomarker and the levels were decreased in cases of individuals who had experienced symptomatic CCM hemorrhage in the prior year ^33^. Furthermore, the CT genotype was associated with low levels of IL-18 and INF-γ (Figure 1), which are increased in patients with epilepsy and CCM hemorrhagic phenotype ^34–36^. The Canonical values derived from the best weighted formula combination with GM-CSF were 2,65X higher in patients who suffered a subsequent symptomatic event (Figure 2). Like this, the a weighted combination of *PTPN2*^rs72872125^ SNP and GM-CSF levels is a potential diagnostic genetic biomarker for the symptomatic phenotype in CCM patients.

We had also provided more evidence that the *FCGR2A^rs1801274^* GG genotype could act as a prognostic biomarker of the disease. The *FCGR2A* gene is located on chromosome 1q23 and consists of seven exons, encoding a member of a family of Fcγ receptors for immunoglobulin G (IgG). Through it is expression in immune system cells such as macrophages, dendritic cells and neutrophils, it is possible to link cellular and humoral immunity^37,38^. The *FCGR2A* rs1801274 variant leads to a substitution of a histidine (A allele) for an arginine (G allele) at position 131, also known as H131R. This polymorphism is capable to increase the binding affinity of FCGR2A to IgG, resulting in activation of the FCGR2A signaling pathway and upregulation of IgG2-dependent phagocytosis ^39,40^. The GG genotype has been associated with several autoimmune diseases, such as Systemic Lupus Erythematosus, Type 1 Diabetes Mellitus, Crohn’s Disease and others, based on its relationship with the release and stimulation of responsive inflammatory processes^41^. Protein structure prediction using bioinformatic tools is an important approach to understand the *FCGR2A* variants. In this study we also performed an *in-silico* analysis that demonstrated that the H131R position is strongly conserved among higher primates, which may be related to its clinical importance.

Lyne et al 2019 showed an upregulation of *FCGR2B* gene in CASH transcriptomic, shedding light on the importance of Fcγ receptors in CCM disease^33^. FcγRII receptors mediates the C-reactive protein (CRP)-induced changes in endothelial function and inflammatory response ^42,43^. A higher binding avidity of CRP to FcγRIIa on immune cells was identified for allotype FcγRIIa-R131 compared to other genotypes ^44^. The variant homozygous genotype (GG) was able to increase the expression of ICAM-1 and E-selectin in HUVEC (Human Umbilical Vein Endothelial Cells) and the levels of tPA, MCP-1, and IL-6 secreted^45^. In addition, the G allele resulted in a significant defect in endothelium dependent vasodilatation and reduced NO activity during endothelial cell stimulation in patients with hypercholesterolaemia, corroborating with what has already been found in models of CCM ^46,47^. This data could potentially impact our comprehension of the pathophysiology of CCM disease as well as consequences with regard to the interpretation of prognostic biomarker for fallow-up of patients.

Growing evidence suggests that vitamin D signaling role in cavernous malformation behavior ^48^. Peripheral plasma vitamin D has been shown to reflect the severity of CCM disease ^49,50^. Indeed, cholecalciferol (vitamin D3), was shown to decrease CCM lesion burden in a murine model of CCM, and to inhibit ROCK activity, known to affect CCM development^22,47^. The effects of vitamin D on the immune system are accomplished by binding to the nuclear Vitamin D Receptor (VDR). Some single nucleotide polymorphisms (SNPs) in genes involved in vitamin D singling were reported to have association with vitamin D deficiency ^51^. The *VDR^rs731236^* also known as TaqI, is a synonymous variant, while *VDR*^rs7975232^ (ApaI) and *VDR^rs^*^11568820^ are located in the 3′ and 5′-untranslated regions of the gene respectively. These SNPs do not alter the amino acid sequence in the VDR protein, but they can exert influence on mRNA stability and gene transcription ^52^. Especially in relation to *VDR*^rs7975232^, the literature does not make clear its role in relation to vitamin D levels, on the other hand we showed that patients who carried the CC genotype have decreased levels of proinflammatory cytokines (Figure 1). Corroborating with a possible protective role of the *VDR*^rs7975232^CC genotype, Jiang et al (2015) showed that A allele slightly increased the risk of temporal lobe epilepsy in children ^53^.

Age at the first symptom is also a fundamental information during the counselling of CCM patients. Distinguishing individuals with higher chances of precocity of symptoms that could influence their productive life is of great importance to the CCM community. Our study provided the correlation between *VDR^rs731236^* AA and *VDR^rs^*^11568820^ CC genotypes to the time to first symptomatic event (Figure 4). However, we could not demonstrate a straight correlation between the *VDR^rs^*^11568820^ CC and *VDR^rs731236^* AA genotype presence and the blood concentration of any of the analyzed cytokines. The relationship between VDR polymorphisms and susceptibility to autoimmunity diseases have been conducted in different settings, while the results obtained so far are conflicting^54–56^. We hypothesized that VDR polymorphism may affect the way proinflammatory cells react to the CCM in an autoimmunity fashion leading to molecular level disruption. To the best of our knowledge, this is the first study to investigate the associations between *VDR* polymorphisms and cerebral cavernous malformations.

Previously reports demonstrated that CCM1/KRIT1 regulates vascular permeability through interaction with CCM2/MGC4607 to stabilize endothelial cell–cell junctions, and together they suppress RhoA activity and, thus, activation of the RhoA effector ROCK^57^. Mutation on these genes lead to augment of actin stress fiber formation and increased permeability of CCMs^58–60^. Moreli et al (2007) have shown that BXL-628, a VDR agonist, prevented RhoA activation and inhibits RhoA/Rho Kinase signaling in rat and human bladder^61^. Since vitamin D is a natural agonist of VDR and animal models have suggested that fasudil and high doses of atorvastatin promotes significant inhibition of CCM lesional development and hemorrhage through the RhoA/Rho Kinase rational, it is reasonable to suppose that the prognostic ability of low levels of serum vitamin D is related to these VD/VDR interactions and its genetic variants.

Our study pioneers a novel classification denoted as the “aggressive behavior” paradigm within cerebral cavernous malformations (CCM), incorporating both hemorrhage and epilepsy as integral components of its symptomatic criteria. This redefined paradigm marks a transformative departure, acknowledging the critical role of epilepsy alongside hemorrhage in defining the clinical spectrum of CCM. Unlike a diagnostic biomarker, which detects the presence of a specific medical condition, a prognostic biomarker assesses the likelihood of progression or stability of medical conditions ^62^. These biomarkers contribute to a more precise and expedited diagnosis, as well as improved patient follow-up. In summary, our study provides new pieces of evidence for possible genetic biomarkers that may influence the behavior of cerebral cavernous malformations. We report a novel observation that the *PTPN2*^rs72872125^ CT and the *VDR*^rs7975232^ CC genotypes were independently associated with an asymptomatic phenotype. Additionally, *PTPN2*^rs72872125^ CC genotype and serum level of GM-CSF could predict a diagnostic association with symptomatic phenotype in CCM patients, while the *FCGR2A^rs1801274^* GG genotype could predict a symptomatic event during follow-up. The study also found a correlation between *VDR*^rs731236^ AA and *VDR*^rs11568820^ CC genotype to the time to first symptomatic event. Overall, this study provided valuable information on the genetic factors that may influence the development and progression of CCM. Moreover, beyond the conventional genetic biomarker purview, our investigation postulates a novel etiological framework. We posit an inflammatory cascade precipitating a self-non-self-interaction, instigating an autoimmune response against CCM. This theoretical construct delineates the changes of CCM from a benign anatomical variant—traditionally construed as a common disease—into an aggressive lesion, now occupying the realm of a rare affliction—the aggressive behavior of CCM. This theoretical construct challenges extant paradigms, accentuating the potential role of immune-mediated mechanisms in the clinical trajectory of CCM, thereby offering prospects for targeted immunomodulatory interventions. In essence, this study augments our comprehension of genetic determinants influencing the advancement of CCM while advancing a transformative hypothesis concerning the disease’s underlying inflammatory dynamics. These insights herald a paradigm shift in conceptualizing the clinical spectrum of CCM, fostering an enriched landscape for future investigative pursuits and potential therapeutic interventions aimed at modulating the intricate inflammatory cascades underpinning the aggressive behavior of CCM.

## Data Availability

All data produced are available online at

## Supplemental material

Table Supplementary 1 (S1)

## Acknowledgements

The authors thank the Cavernoma Alliance Brazil Research Institute – Aliança Cavernoma Brasil for the logistic assistance.

## Sources of Funding

The author(s) disclosed receipt of the following financial support for the research, authorship, and/or publication of this article: Brazilian National Council for Scientific and Technological Development (CNPq Number 440779/2016-2), Senator Romario Faria parliamentary amendment (no. 37990007 EIND), financial support of Coordination for the Improvement of Higher Education Personnel (CAPES Number 88887.130752/2016-00), FAPERJ E-26/210.657/2021, E-26/210.273/2018 and E-26/201.040/2021 and Chamada Pública MCTI/FINEP/CT-INFRA-PROINFRA 02/2014 – Equipamentos Multiusuários – Ref. n° 0097/2016. Thanks to Fundação de Amparo à Pesquisa do Estado do Rio de Janeiro (FAPERJ) and Casa Hunter for the financial help to this Project.

## Disclosures

The authors declare no competing interests

## Notes

### Competing Interest Statement

The authors have declared no competing interest.

### Funding Statement

This study was funded by CNPQ, CAPES, FAPERJ, Senator Romario Faria, MS and Casa Hunter

### Author Declarations

Qualified researcher on human subjects conducted this study having been approved by the National Council for Ethics in Research (CAAE 69409617.9.0000.5258).

## References

1. Flemming KD, Graff-Radford J, Aakre J, Kantarci K, Lanzino G, Brown RDJ, Mielke MM, Roberts RO, Kremers W, Knopman DS, et al. Population-Based Prevalence of Cerebral Cavernous Malformations in Older Adults: Mayo Clinic Study of Aging. JAMA Neurol. 2017;74:801– 805.

2. Spiegler S, Rath M, Paperlein C, Felbor U. Cerebral Cavernous Malformations: An Update on Prevalence, Molecular Genetic Analyses, and Genetic Counselling. Mol. Syndromol. 2018;9:60–69.

3. de Souza JM, Domingues RC, Cruz LCH, Domingues FS, Iasbeck T, Gasparetto EL. Susceptibility-Weighted Imaging for the Evaluation of Patients with Familial Cerebral Cavernous Malformations: A Comparison with T2-Weighted Fast Spin-Echo and Gradient-Echo Sequences. Am. J. Neuroradiol. [Internet]. 2008;29:154–158. Available from: http://www.ajnr.org/lookup/doi/10.3174/ajnr.A0748

4. Salman RA-S, Hall JM, Horne MA, Moultrie F, Josephson CB, Bhattacharya JJ, Counsell CE, Murray GD, Papanastassiou V, Ritchie V, et al. Untreated clinical course of cerebral cavernous malformations: a prospective, population-based cohort study. Lancet Neurol. [Internet]. 2012;11:217–224. Available from: 10.1016/S1474-4422(12)70004-2

5. Ironside JW. Pathology of tumours of the nervous system. (5th ed.). D. S. Russell and L. J. Rubinstein. Edward Arnold, London, 1989. No. of pages: 1012. Price: £110.00. ISBN: 07131 4549 8. J. Pathol. [Internet]. 1989;158:359–359. Available from: 10.1002/path.1711580413

6. Weinsheimer S, Nelson J, Abla AA, Ko NU, Tsang C, Okoye O, Zabramski JM, Akers A, Zafar A, Mabray MC, et al. Intracranial Hemorrhage Rate and Lesion Burden in Patients With Familial Cerebral Cavernous Malformation. J. Am. Heart Assoc. [Internet]. 2023;12:e027572. Available from: https://www.ahajournals.org/doi/10.1161/JAHA.122.027572

7. Dashti SR, Hoffer A, Hu YC, Selman WR. Molecular genetics of familial cerebral cavernous malformations. Neurosurg. Focus. 2006;21:e2.

8. Gross BA, Lin N, Du R, Day AL. The natural history of intracranial cavernous malformations. Neurosurg. Focus. 2011;30:E24.

9. Labauge P, Denier C, Bergametti F, Tournier-Lasserve E. Genetics of cavernous angiomas. Lancet Neurol. [Internet]. 2007;6:237–244. Available from: https://linkinghub.elsevier.com/retrieve/pii/S1474442207700534

10. Gault J, Sain S, Hu L-J, Awad IA. SPECTRUM OF GENOTYPE AND CLINICAL MANIFESTATIONS IN CEREBRAL CAVERNOUS MALFORMATIONS. Neurosurgery [Internet]. 2006;59:1278–1285. Available from: https://journals.lww.com/00006123-200612000-00015

11. Snellings DA, Hong CC, Ren AA, Lopez-Ramirez MA, Girard R, Srinath A, Marchuk DA, Ginsberg MH, Awad IA, Kahn ML. Cerebral Cavernous Malformation: From Mechanism to Therapy. Circ. Res. [Internet]. 2021;129:195–215. Available from: https://www.ahajournals.org/doi/10.1161/CIRCRESAHA.121.318174

12. Koskimäki J, Polster SP, Li Y, Romanos S, Srinath A, Zhang D, Carrión-Penagos J, Lightle R, Moore T, Lyne SB, et al. Common transcriptome, plasma molecules, and imaging signatures in the aging brain and a Mendelian neurovascular disease, cerebral cavernous malformation. GeroScience [Internet]. 2020;42:1351–1363. Available from: https://link.springer.com/10.1007/s11357-020-00201-4

13. Shenkar R, Shi C, Check IJ, Lipton HL, Awad IA. Concepts and hypotheses: inflammatory hypothesis in the pathogenesis of cerebral cavernous malformations. Neurosurgery. 2007;61:693.

14. Shi C, Shenkar R, Du H, Duckworth E, Raja H, Batjer HH, Awad IA. Immune response in human cerebral cavernous malformations. Stroke. 2009;40:1659–1665.

15. Shi C, Shenkar R, Kinloch A, Henderson SG, Shaaya M, Chong AS, Clark MR, Awad IA. Immune complex formation and in situ B-cell clonal expansion in human cerebral cavernous malformations. J. Neuroimmunol. 2014;272:67–75.

16. Plummer NW, Gallione CJ, Srinivasan S, Zawistowski JS, Louis DN, Marchuk DA. Loss of p53 sensitizes mice with a mutation in Ccm1 (KRIT1) to development of cerebral vascular malformations. Am. J. Pathol. 2004;165:1509–1518.

17. Maddaluno L, Rudini N, Cuttano R, Bravi L, Giampietro C, Corada M, Ferrarini L, Orsenigo F, Papa E, Boulday G, et al. EndMT contributes to the onset and progression of cerebral cavernous malformations. Nature. 2013;498:492–496.

18. Fontes-Dantas FL, da Fontoura Galvão G, Veloso da Silva E, Alves-Leon S, Cecília da Silva Rêgo C, Garcia DG, Marques SA, Blanco Martinez AM, Reis da Silva M, Marcondes de Souza J. Novel CCM1 (KRIT1) Mutation Detection in Brazilian Familial Cerebral Cavernous Malformation: Different Genetic Variants in Inflammation, Oxidative Stress, and Drug Metabolism Genes Affect Disease Aggressiveness. World Neurosurg. 2020;138:535–540.e8.

19. Choquet H, Pawlikowska L, Nelson J, McCulloch CE, Akers A, Baca B, Khan Y, Hart B, Morrison L, Kim H. Polymorphisms in Inflammatory and Immune Response Genes Associated with Cerebral Cavernous Malformation Type 1 Severity. Cerebrovasc. Dis. [Internet]. 2014;38:433–440. Available from: https://www.karger.com/Article/FullText/369200

20. Tang AT, Choi JP, Kotzin JJ, Yang Y, Hong CC, Hobson N, Girard R, Zeineddine HA, Lightle R, Moore T, et al. Endothelial TLR4 and the microbiome drive cerebral cavernous malformations. Nature. 2017;545:305–310.

21. Perrelli A, Retta SF. Polymorphisms in genes related to oxidative stress and inflammation: Emerging links with the pathogenesis and severity of Cerebral Cavernous Malformation disease. Free Radic. Biol. Med. [Internet]. 2021;172:403–417. Available from: https://linkinghub.elsevier.com/retrieve/pii/S0891584921003865

22. da Fontoura Galváo G, Fontes-Dantas FL, da Silva EV, Alves-Leon SV, de Souza JM. Association of Variants in FCGR2A, PTPN2, and GM-CSF with Cerebral Cavernous Malformation: Potential Biomarkers for a Symptomatic Disease. Curr. Neurovasc. Res. [Internet]. 2021;18:172–180. Available from: https://www.eurekaselect.com/193838/article

23. Lai CC, Nelsen B, Frias-Anaya E, Gallego-Gutierrez H, Orecchioni M, Herrera V, Ortiz E, Sun H, Mesarwi OA, Ley K, et al. Neuroinflammation Plays a Critical Role in Cerebral Cavernous Malformation Disease. Circ. Res. [Internet]. 2022;131:909–925. Available from: https://www.ahajournals.org/doi/10.1161/CIRCRESAHA.122.321129

24. McCole DF. Regulation of epithelial barrier function by the inflammatory bowel disease candidate gene, PTPN2. Ann. N. Y. Acad. Sci. [Internet]. 2012;1257:108–114. Available from: https://onlinelibrary.wiley.com/doi/10.1111/j.1749-6632.2012.06522.x

25. Bottini N, Peterson EJ. Tyrosine Phosphatase PTPN22: Multifunctional Regulator of Immune Signaling, Development, and Disease. Annu. Rev. Immunol. [Internet]. 2014;32:83–119. Available from: https://www.annualreviews.org/doi/10.1146/annurev-immunol-032713-120249

26. Santin I, Moore F, Colli ML, Gurzov EN, Marselli L, Marchetti P, Eizirik DL. PTPN2, a Candidate Gene for Type 1 Diabetes, Modulates Pancreatic β-Cell Apoptosis via Regulation of the BH3-Only Protein Bim. Diabetes [Internet]. 2011;60:3279–3288. Available from: https://diabetesjournals.org/diabetes/article/60/12/3279/14490/PTPN2-a-Candidate-Gene-for-Type-1-Diabetes

27. Hsieh W-C, Svensson MN, Zoccheddu M, Tremblay ML, Sakaguchi S, Stanford SM, Bottini N. PTPN2 links colonic and joint inflammation in experimental autoimmune arthritis. JCI Insight [Internet]. 2020;5. Available from: https://insight.jci.org/articles/view/141868

28. Shaw AM, Qasem A, Naser SA. Modulation of PTPN2/22 Function by Spermidine in CRISPR-Cas9-Edited T-Cells Associated with Crohn’s Disease and Rheumatoid Arthritis. Int. J. Mol. Sci. [Internet]. 2021;22:8883. Available from: https://www.mdpi.com/1422-0067/22/16/8883

29. Ramagopalan S V, Heger A, Berlanga AJ, Maugeri NJ, Lincoln MR, Burrell A, Handunnetthi L, Handel AE, Disanto G, Orton S-M, et al. A ChIP-seq defined genome-wide map of vitamin D receptor binding: Associations with disease and evolution. Genome Res. [Internet]. 2010;20:1352–1360. Available from: http://genome.cshlp.org/lookup/doi/10.1101/gr.107920.110

30. Ellis JA, Scurrah KJ, Li YR, Ponsonby A-L, Chavez RA, Pezic A, Dwyer T, Akikusa JD, Allen RC, Becker ML, et al. Epistasis amongst PTPN2 and genes of the vitamin D pathway contributes to risk of juvenile idiopathic arthritis. J. Steroid Biochem. Mol. Biol. [Internet]. 2015;145:113–120. Available from: https://linkinghub.elsevier.com/retrieve/pii/S0960076014002477

31. Murray PJ. The primary mechanism of the IL-10-regulated antiinflammatory response is to selectively inhibit transcription. Proc. Natl. Acad. Sci. [Internet]. 2005;102:8686–8691. Available from: https://pnas.org/doi/full/10.1073/pnas.0500419102

32. Sabat R, Grütz G, Warszawska K, Kirsch S, Witte E, Wolk K, Geginat J. Biology of interleukin-10. Cytokine Growth Factor Rev. [Internet]. 2010;21:331–344. Available from: https://linkinghub.elsevier.com/retrieve/pii/S1359610110000651

33. Lyne SB, Girard R, Koskimäki J, Zeineddine HA, Zhang D, Cao Y, Li Y, Stadnik A, Moore T, Lightle R, et al. Biomarkers of cavernous angioma with symptomatic hemorrhage. JCI Insight [Internet]. 2019;4. Available from: https://insight.jci.org/articles/view/128577

34. Mochol M, Taubøll E, Aukrust P, Ueland T, Andreassen OA, Svalheim S. Interleukin 18 (IL-18) and its binding protein (IL-18BP) are increased in patients with epilepsy suggesting low-grade systemic inflammation. Seizure [Internet]. 2020;80:221–225. Available from: https://linkinghub.elsevier.com/retrieve/pii/S1059131120301473

35. Gao F, Gao Y, Zhang S -j., Zhe X, Meng F-L, Qian H, Zhang B, Li Y-J. Alteration of plasma cytokines in patients with active epilepsy. Acta Neurol. Scand. [Internet]. 2017;135:663–669. Available from: https://onlinelibrary.wiley.com/doi/10.1111/ane.12665

36. Girard R, Zeineddine HA, Fam MD, Mayampurath A, Cao Y, Shi C, Shenkar R, Polster SP, Jesselson M, Duggan R, et al. Plasma Biomarkers of Inflammation Reflect Seizures and Hemorrhagic Activity of Cerebral Cavernous Malformations. Transl. Stroke Res. [Internet]. 2018;9:34–43. Available from: http://link.springer.com/10.1007/s12975-017-0561-3

37. Zhang C, Wang W, Zhang H, Wei L, Guo S. Association of FCGR2A rs1801274 polymorphism with susceptibility to autoimmune diseases: A meta-analysis. Oncotarget. 2016;7:39436–39443.

38. Duan J, Lou J, Zhang Q, Ke J, Qi Y, Shen N, Zhu B, Zhong R, Wang Z, Liu L, et al. A genetic variant rs1801274 in FCGR2A as a potential risk marker for Kawasaki disease: a case-control study and meta-analysis. PLoS One. 2014;9:e103329.

39. Warmerdam PA, van de Winkel JG, Vlug A, Westerdaal NA, Capel PJ. A single amino acid in the second Ig-like domain of the human Fc gamma receptor II is critical for human IgG2 binding. J. Immunol. [Internet]. 1991;147:1338–1343. Available from: https://journals.aai.org/jimmunol/article/147/4/1338/24700/A-single-amino-acid-in-the-second-Ig-like-domain

40. Shrestha S, Wiener HW, Olson AK, Edberg JC, Bowles NE, Patel H, Portman MA. Functional FCGR2B gene variants influence intravenous immunoglobulin response in patients with Kawasaki disease. J. Allergy Clin. Immunol. [Internet]. 2011;128:677–680.e1. Available from: https://linkinghub.elsevier.com/retrieve/pii/S0091674911006646

41. Hayat S, Babu G, Das A, Howlader ZH, Mahmud I, Islam Z. Fc-gamma IIIa-V158F receptor polymorphism contributes to the severity of Guillain-Barré syndrome. Ann. Clin. Transl. Neurol. 2020;7:1040–1049.

42. Mineo C, Gormley AK, Yuhanna IS, Osborne-Lawrence S, Gibson LL, Hahner L, Shohet R V, Black S, Salmon JE, Samols D, et al. FcγRIIB Mediates C-Reactive Protein Inhibition of Endothelial NO Synthase. Circ. Res. [Internet]. 2005;97:1124–1131. Available from: https://www.ahajournals.org/doi/10.1161/01.RES.0000194323.77203.fe

43. Ryu J, Lee C, Shin J, Park C, Kim J, Park S, Han K. FcγRIIa mediates C-reactive protein-induced inflammatory responses of human vascular smooth muscle cells by activating NADPH oxidase 4. Cardiovasc. Res. [Internet]. 2007;75:555–565. Available from: 10.1016/j.cardiores.2007.04.027

44. Stein M-P, Edberg JC, Kimberly RP, Mangan EK, Bharadwaj D, Mold C, Du Clos TW. C-reactive protein binding to FcγRIIa on human monocytes and neutrophils is allele-specific. J. Clin. Invest. [Internet]. 2000;105:369–376. Available from: http://www.jci.org/articles/view/7817

45. Raaz-Schrauder D, Ekici AB, Klinghammer L, Stumpf C, Achenbach S, Herrmann M, Reis A, Garlichs CD. The proinflammatory effect of C-reactive protein on human endothelial cells depends on the FcγRIIa genotype. Thromb. Res. [Internet]. 2014;133:426–432. Available from: https://linkinghub.elsevier.com/retrieve/pii/S0049384813006166

46. Schneider MP, Leusen JHW, Herrmann M, Garlichs CD, Amann K, John S, Schmieder RE. The Fcγ receptor IIA R131H gene polymorphism is associated with endothelial function in patients with hypercholesterolaemia. Atherosclerosis [Internet]. 2011;218:411–415. Available from: https://linkinghub.elsevier.com/retrieve/pii/S0021915011005880

47. Gibson CC, Zhu W, Davis CT, Bowman-Kirigin JA, Chan AC, Ling J, Walker AE, Goitre L, Delle Monache S, Retta SF, et al. Strategy for Identifying Repurposed Drugs for the Treatment of Cerebral Cavernous Malformation. Circulation [Internet]. 2015;131:289–299. Available from: https://www.ahajournals.org/doi/10.1161/CIRCULATIONAHA.114.010403

48. Awad IA, Polster SP. Cavernous angiomas: deconstructing a neurosurgical disease. J. Neurosurg. 2019;131:1–13.

49. Girard R, Khanna O, Shenkar R, Zhang L, Wu M, Jesselson M, Zeineddine HA, Gangal A, Fam MD, Gibson CC, et al. Peripheral plasma vitamin D and non-HDL cholesterol reflect the severity of cerebral cavernous malformation disease. Biomark. Med. 2016;10:255–264.

50. Akers A, Al-Shahi Salman R, A. Awad I, Dahlem K, Flemming K, Hart B, Kim H, Jusue-Torres I, Kondziolka D, Lee C, et al. Synopsis of Guidelines for the Clinical Management of Cerebral Cavernous Malformations: Consensus Recommendations Based on Systematic Literature Review by the Angioma Alliance Scientific Advisory Board Clinical Experts Panel. Neurosurgery [Internet]. 2017;80:665–680. Available from: https://journals.lww.com/00006123-201705000-00012

51. Krasniqi E, Boshnjaku A, Wagner K-H, Wessner B. Association between Polymorphisms in Vitamin D Pathway-Related Genes, Vitamin D Status, Muscle Mass and Function: A Systematic Review. Nutrients [Internet]. 2021;13:3109. Available from: https://www.mdpi.com/2072-6643/13/9/3109

52. Uitterlinden AG, Fang Y, van Meurs JBJ, Pols HAP, van Leeuwen JPTM. Genetics and biology of vitamin D receptor polymorphisms. Gene [Internet]. 2004;338:143–156. Available from: https://linkinghub.elsevier.com/retrieve/pii/S0378111904003075

53. Jiang P, Zhu W-Y, He X, Tang M-M, Dang R-L, Li H-D, Xue Y, Zhang L-H, Wu Y-Q, Cao L-J. Association between Vitamin D Receptor Gene Polymorphisms with Childhood Temporal Lobe Epilepsy. Int. J. Environ. Res. Public Health [Internet]. 2015;12:13913–13922. Available from: http://www.mdpi.com/1660-4601/12/11/13913

54. Fouad H, Yahia S, Elsaid A, Hammad A, Wahba Y, El-Gilany A-H, Abdel-Aziz A-AF. Oxidative stress and vitamin D receptor BsmI gene polymorphism in Egyptian children with systemic lupus erythematosus: a single center study. Lupus. 2019;28:771–777.

55. Ghaly MS, Badra DI, Dessouki O, Elmaraghy NN, Hassan R. Vitamin D receptor Fok1 & Bsm 1 Gene Polymorphisms in Systemic Lupus Erythematosus and Osteoarthritis: Autoimmune Inflammatory versus Degenerative Model. Egypt. J. Immunol. 2017;24:151–164.

56. Tizaoui K, Hamzaoui K. Association between VDR polymorphisms and rheumatoid arthritis disease: Systematic review and updated meta-analysis of case-control studies. Immunobiology. 2015;220:807–816.

57. Stockton RA, Shenkar R, Awad IA, Ginsberg MH. Cerebral cavernous malformations proteins inhibit Rho kinase to stabilize vascular integrity. J. Exp. Med. 2010;207:881–896.

58. Borikova AL, Dibble CF, Sciaky N, Welch CM, Abell AN, Bencharit S, Johnson GL. Rho kinase inhibition rescues the endothelial cell cerebral cavernous malformation phenotype. J. Biol. Chem. 2010;285:11760– 11764.

59. Whitehead KJ, Chan AC, Navankasattusas S, Koh W, London NR, Ling J, Mayo AH, Drakos SG, Jones CA, Zhu W, et al. The cerebral cavernous malformation signaling pathway promotes vascular integrity via Rho GTPases. Nat. Med. 2009;15:177–184.

60. Crose LES, Hilder TL, Sciaky N, Johnson GL. Cerebral cavernous malformation 2 protein promotes smad ubiquitin regulatory factor 1-mediated RhoA degradation in endothelial cells. J. Biol. Chem. 2009;284:13301–13305.

61. Morelli A, Vignozzi L, Filippi S, Vannelli GB, Ambrosini S, Mancina R, Crescioli C, Donati S, Fibbi B, Colli E, et al. BXL-628, a vitamin D receptor agonist effective in benign prostatic hyperplasia treatment, prevents RhoA activation and inhibits RhoA/Rho kinase signaling in rat and human bladder. Prostate. 2007;67:234–247.

62. Amur S, LaVange L, Zineh I, Buckman-Garner S, Woodcock J. Biomarker Qualification: Toward a Multiple Stakeholder Framework for Biomarker Development, Regulatory Acceptance, and Utilization. Clin. Pharmacol. Ther. [Internet]. 2015;98:34–46. Available from: https://onlinelibrary.wiley.com/doi/10.1002/cpt.136

63. von Elm E, Altman DG, Egger M, Pocock SJ, Gøtzsche PC, Vandenbroucke JP. The Strengthening the Reporting of Observational Studies in Epidemiology (STROBE) statement: guidelines for reporting observational studies. Lancet [Internet]. 2007;370:1453–1457. Available from: https://linkinghub.elsevier.com/retrieve/pii/S014067360761602X

64. Fontes FL, de Araújo LF, Coutinho LG, Leib SL, Agnez-Lima LF. Genetic polymorphisms associated with the inflammatory response in bacterial meningitis. BMC Med. Genet. [Internet]. 2015;16:1–12. Available from: 10.1186/s12881-015-0218-6

65. Chehuen Bicalho V, da Fontoura Galvão G, Lima Fontes-Dantas F, Paulo da Costa Gonçalves J, Dutra de Araujo A, Carolina França L, Emílio Corrêa Leite P, Campolina Vidal D, Castro Filho R, Vieira Alves-Leon S, et al. Asymptomatic cerebral cavernous angiomas associated with plasma marker signature. J. Clin. Neurosci. [Internet]. 2021;89:258–263. Available from: https://linkinghub.elsevier.com/retrieve/pii/S0967586821001910

66. da Fontoura Galvão G, da Silva EV, Trefilio LM, Alves-Leon SV, Fontes-Dantas FL, de Souza JM. Comprehensive CCM3 Mutational Analysis in Two Patients with Syndromic Cerebral Cavernous Malformation. Transl. Stroke Res. [Internet]. 2023;Available from: https://link.springer.com/10.1007/s12975-023-01131-x

67. Pettersen EF, Goddard TD, Huang CC, Couch GS, Greenblatt DM, Meng EC, Ferrin TE. UCSF Chimera?A visualization system for exploratory research and analysis. J. Comput. Chem. [Internet]. 2004;25:1605–1612. Available from: https://onlinelibrary.wiley.com/doi/10.1002/jcc.20084

68. Edgar RC. MUSCLE: multiple sequence alignment with high accuracy and high throughput. Nucleic Acids Res. [Internet]. 2004;32:1792–1797. Available from: https://academic.oup.com/nar/article-lookup/doi/10.1093/nar/gkh340

